# Tie2 activation protects against prothrombotic endothelial dysfunction in COVID-19

**DOI:** 10.1101/2021.05.13.21257070

**Authors:** Alec A. Schmaier, Gabriel Pajares Hurtado, Zachary J. Manickas-Hill, Kelsey D. Sack, Siyu M Chen, Victoria Bhambhani, Juweria Quadir, Anjali K. Nath, Ai-ris Y. Collier, Debby Ngo, Dan H. Barouch, Robert E. Gerszten, Xu G. Yu, MGH COVID-19 Collection and Processing Team, Kevin Peters, Robert Flaumenhaft, Samir M. Parikh

## Abstract

Profound endothelial dysfunction accompanies the microvascular thrombosis commonly observed in severe COVID-19. In the quiescent state, the endothelial surface is anticoagulant, a property maintained at least in part via constitutive signaling through the Tie2 receptor. During inflammation, the Tie2 antagonist angiopoietin-2 (Angpt-2) is released from activated endothelial cells and inhibits Tie2, promoting a prothrombotic phenotypic shift. We sought to assess whether severe COVID-19 is associated with procoagulant dysfunction of the endothelium and alterations in the Tie2-angiopoietin axis. Primary human endothelial cells treated with plasma from patients with severe COVID-19 upregulated the expression of thromboinflammatory genes, inhibited expression of antithrombotic genes, and promoted coagulation on the endothelial surface. Pharmacologic activation of Tie2 with the small molecule AKB-9778 reversed the prothrombotic state induced by COVID-19 plasma in primary endothelial cells. On lung autopsy specimens from COVID-19 patients, we found a prothrombotic endothelial signature as evidenced by increased von Willebrand Factor and loss of anticoagulant proteins. Assessment of circulating endothelial markers in a cohort of 98 patients with mild, moderate, or severe COVID-19 revealed profound endothelial dysfunction indicative of a prothrombotic state. Angpt-2 concentrations rose with increasing disease severity and highest levels were associated with worse survival. These data highlight the disruption of Tie2-angiopoietin signaling and procoagulant changes in endothelial cells in severe COVID-19. Moreover, our findings provide novel rationale for current trials of Tie2 activating therapy with AKB-9778 in severe COVID-19 disease.

## Introduction

Severe acute respiratory syndrome coronavirus 2 (SARS-CoV-2), the virus responsible for coronavirus disease (COVID-19), can result in critical illness characterized by severe pulmonary manifestations^1^, in addition to several extrapulmonary manifestations^2^, and carries a significant morality rate. Critical COVID-19 illness is characterized by a prothrombotic coagulopathy, and higher D-dimer concentrations and activation of coagulation are associated with worse outcomes^3–5^. Fibrin deposition in the lung vasculature is a commonly identified histopathologic finding, and microvascular thrombosis may be a key driver of COVID-19 pathophysiology^6,7^. Multiple lines of evidence, including measurement of circulating proteins and metabolites and analysis of histologic specimens, have demonstrated severe vascular inflammation and endothelial injury^8^. Therefore, a phenotypic switch of endothelial cells to a procoagulant state appears to be a critical disease mechanism. Endothelial dysfunction in COVID-19 may be mediated through circulating inflammatory cytokines^9,10^, auto-antibodies^11,12^, neutrophil extracellular traps (NETs)^13^, and potentially via direct viral infection^14^.

Procoagulant changes in endothelial cells can be characterized by loss of constitutive anticoagulant function and/or upregulation of thromboinflammatory mediators. Several studies have demonstrated that severe COVID-19 is associated with increased circulating levels of such markers, including procoagulant Von Willebrand factor (VWF)^15^ and plasminogen activator inhibitor (PAI-1)^16^, vascular cell adhesion markers (VCAM, ICAM, E-selectin)^17–19^, and increased circulating antithrombotic endothelial surface proteins tissue factor pathway inhibitor (TFPI)^20^ and thrombomodulin^21^, which are cleaved off the endothelial surface during inflammation^22^.

We have previously demonstrated via unbiased proteomics that endothelial dysfunction is strongly implicated in the coagulopathy of critical illness^23^. Specifically, dysregulation of the Tie2-angiopoietin pathway emerged as a central link between vascular inflammation and inappropriate coagulation^23^. The receptor tyrosine kinase Tie2 is highly enriched in vascular endothelium, and its ligand angiopoietin 1 (Angpt-1) promotes vascular stability and quiescence through Tie2 activation^24^. Moreover, activation of the Tie2 pathway can prevent the heightened thrombosis that occurs during septic endothelial injury^23^. Angiopoietin 2 (Angpt-2) is an Angpt-1 paralog that competitively inhibits Tie2. Angpt-2 expression is promoted by tissue hypoxia and other inflammatory pathways and is subsequently stored in endothelial Weibel-Palade bodies and released during endothelial activation^25^. Angpt-2 levels are even further elevated in the plasma of septic patients with disseminated intravascular coagulation (DIC) compared to those with sepsis but without DIC, and high Angpt-2 levels potentiate endothelial dysfunction of critical illness^23,24^. Loss of constitutive Tie2 signaling is mediated through Angpt-2 antagonism or by Tie2 cleavage from the endothelial surface^26,27^. Inhibition of Tie2 results in loss of barrier function and anti-inflammatory transcriptional machinery, most notably characterized by upregulation of Angpt-2 itself, leading to a positive feedback loop and further Tie2 suppression. Tie2 kinase activity is also tonically inhibited by vascular endothelial protein tyrosine phosphatase (VE-PTP). Similar to Angpt-2, VE-PTP expression is also increased under conditions of endothelial stress such as hypoxia, creating an Angpt-1 resistant state^28^. A first-in-class VE-PTP inhibitor small molecule, AKB-9778 (Razuprotafib) restores cytoprotective Tie2 signaling to protect EC function in the presence of inflammatory stimuli^28–30^.

Since Angpt-2 contributes to the coagulopathy of sepsis^23^, we hypothesized that Angpt-2 may also regulate the host endothelial response in COVID-19 and, more importantly, represent a therapeutic target. We used a cell culture model to perturb theTie2-angiopoietin system and determine if increasing COVID-19 disease severity leads to procoagulant changes in the endothelium. We found that plasma from COVID-19 patients can directly promote a prothrombotic state in endothelial cells and that pharmacologic activation of Tie2 signaling rescues endothelial antithrombotic function. We established the presence of prothrombotic endothelial markers in lung tissue biopsies from COVID-19 patients. We next validated the relationship between COVID-19 severity and endothelial dysfunction by measuring Angpt-2 and other markers of thromboinflammatory endothelial activation in plasma from patients with mild, moderate and severe COVID-19. Markers of endothelial dysfunction and thrombosis are strongly correlated with COVID-19 disease severity and survival. Our findings indicate a role for Angpt-2-Tie2 system in endothelial dysfunction in COVID-19 pathogenesis and offer potential targets for therapeutic intervention.

## Results

### COVID-19 Cohort

Plasma was collected from 98 patients with PCR-confirmed SARS-CoV-2 infection: 42 were admitted to an intensive care unit (ICU), 37 were hospitalized in a non-ICU medical floor, and 19 were outpatients. In addition, we studied 12 historical healthy controls. Patient characteristics are shown in **Table 1**. The median time from symptom onset to sample collection was 12 days (interquartile range 7-20 days). Patients with COVID-19 requiring ICU admission, compared to hospitalized patients not requiring ICU-level care, were more often male, but otherwise similar in terms of demographics and clinical history. History of obesity, cardiovascular disease, or diabetes were not associated with COVID-19 illness severity among hospitalized patients. The majority of ICU patients required mechanical ventilation (38, 90.5 %) and experienced a much longer duration of hospitalization than non-ICU patients (27.5 versus 6 median days, *P* < 0.0001). Twelve (28.5%) ICU patients died during their index hospitalization whereas all non-ICU patients (*n* = 37) survived until hospital discharge.

**Table 1.** Demographic, clinical, and outcome data of patients. BMI, body mass index; COPD, chronic obstructive pulmonary disease; IQR, interquartile range; NA, not available; VTE, venous thromboembolism.

|  | Hospitalized<br>(n = 79) | ICU<br>(n = 42) | non-ICU<br>(n = 37) | Outpatients<br>(n = 19) | Healthy<br>Controls<br>(n = 12) | P value<br>(ICU vs<br>non-ICU) |
| --- | --- | --- | --- | --- | --- | --- |
| Age – years median<br>(IQR) | 63 (52-77) | 66 (53-74) | 60 (41-80) | 50 (32-57) | 31.5 (24-59) | 0.44 |
| Male sex - n (%) | 48 (60.8) | 32 (76.2) | 16 (43.2) | 5 (26.3) | 5 (41.7) | 0.015 |
| BMI – kg/m<sup>2</sup> median<br>(IQR) | 27.4 (25.6-31.9) | 27.8 (24.0-52.2) | 25.7 (24.0-51.0) | 32.0 (24.9-45.2) | 24.6 (21.1-30.7) | 0.65 |
| **Ethnicity - n (%)** |  |  |  |  |  |  |
| White | 29 (36.7) | 17 (40.4) | 12 (32.4) | 15 (78.9) | 10 (83.3) | 0.19 |
| Black | 19 (27.6) | 11 (26.1) | 8 (21.6) | 3 (15.8) | 0 |  |
| Hispanic | 23 (29.1) | 10 (23.8) | 13 (35.1) | 0 | 0 |  |
| Asian | 3 (3.8) | 0 | 3 (8.1) | 0 | 1 (8.3) |  |
| Other / NA | 5 (6.3) | 4 (9.5) | 1 (2.7) | 1 (5.3) | 1 (8.3) |  |
| **Comorbidities** |  |  |  |  |  |  |
| Obesity - n (%) | 32 (40.5) | 17 (40.4) | 15 (40.5) | 10 (52.6) | 1 (8.3) | 0.99 |
| Hypertension - n (%) | 39 (49.4) | 20 (47.6) | 19 (51.3) | 4 (21.0) | 1 (8.3) | 0.74 |
| Hyperlipidemia – n<br>(%) | 21 (26.6) | 9 (21.4) | 14 (37.8) | 3 (15.8) | 1 (8.3) | 0.11 |
| Coronary artery<br>disease – n (%) | 10 (12.7) | 6 (14.3) | 4 (10.8) | 1 (5.2) | 0 | 0.85 |
| Congestive heart<br>failure – n (%) | 8 (10.1) | 3 (7.1) | 5 (13.5) | 0 | 0 | 0.57 |
| History of smoking –<br>n (%) | 23 (29.1) | 12 (28.6) | 11 (29.7) | 5 (13.5) | 0 | 0.91 |
| COPD or asthma – n<br>(%) | 17 (21.5) | 7 (16.7) | 10 (27) | 5 (13.5) | 0 | 0.26 |
| Diabetes – n (%) | 30 (40.0) | 17 (40.4) | 13 (35.1) | 2 (10.5) | 0 | 0.63 |
| History of stroke | 4 (5.1) | 1 (2.4) | 3 (8.1) | 0 | 0 | 0.25 |
| Chronic or end-stage renal disease<br>– n (%) | 10 (12.7) | 8 (19.0) | 2 (2.7) | 0 | 0 | 0.07 |
| **Outcomes** |  |  |  |  |  |  |
| Mechanical<br>ventilation – n (%) | 38 (48.1) | 38 (90.5) | 0 | - | - | <0.0001 |
| VTE or catheter<br>thrombosis – n (%) | 15 (19.0) | 10 (23.8) | 5 (13.5) | - | - | 0.24 |
| Acute kidney injury –<br>n (%) | 39 (49.4) | 31 (73.8) | 8 (21.6) | - | - | <0.0001 |
| In-hospital mortality<br>– n (%) | 12 (15.2) | 12 (28.5) | 0 | - | - | <0.0001 |
| Length of<br>hospitalization,<br>median days – n (%) | 16 | 27.5 | 6 | - | - | <0.0001 |

### Endothelial cells treated with plasma from COVID-19 patients exhibit upregulation of thromboinflammatory genes and promotion of coagulation on the endothelial surface, which is rescued by Tie2 activation

Plasma from patients with severe COVID-19 contains a wide array of pro-inflammatory factors including cytokines^9,10^, auto-antibodies^11,12^, cell-free DNA^31^, and NETs^13^. To determine if the plasma milieu is capable of inducing procoagulant changes in the endothelium, we cultured human umbilical vein endothelial cells (HUVECs) in the presence 10% plasma from patients with severe (ICU), moderate (non-ICU), mild (outpatient) COVID-19, or from healthy controls. Plasma from patients with severe or moderate COVID-19 induced significant upregulation of tissue factor, E-selectin, and Angpt-2 gene expression and concurrently decreased expression of antithrombotic genes EPCR, TFPI, and thrombomodulin (**Figure 1**). Plasma from patients with severe COVID-19 did not significantly alter Tie2 or VE-PTP gene expression (**Figure 1**) Given the significant induction of Angpt-2 and its association with the coagulopathy of critical illness via inhibition of Tie2^23^, we tested whether pharmacologic activation of Tie2 could reverse thromboinflammatory gene expression triggered by COVID-19 plasma. Indeed, stimulation of Tie2 by recombinant Angpt-1 or the small molecule VE-PTP inhibitor AKB-9778 normalized much of the gene expression changes induced by plasma from COVID-19 patients (**Figure 1**).

**Figure 1.**
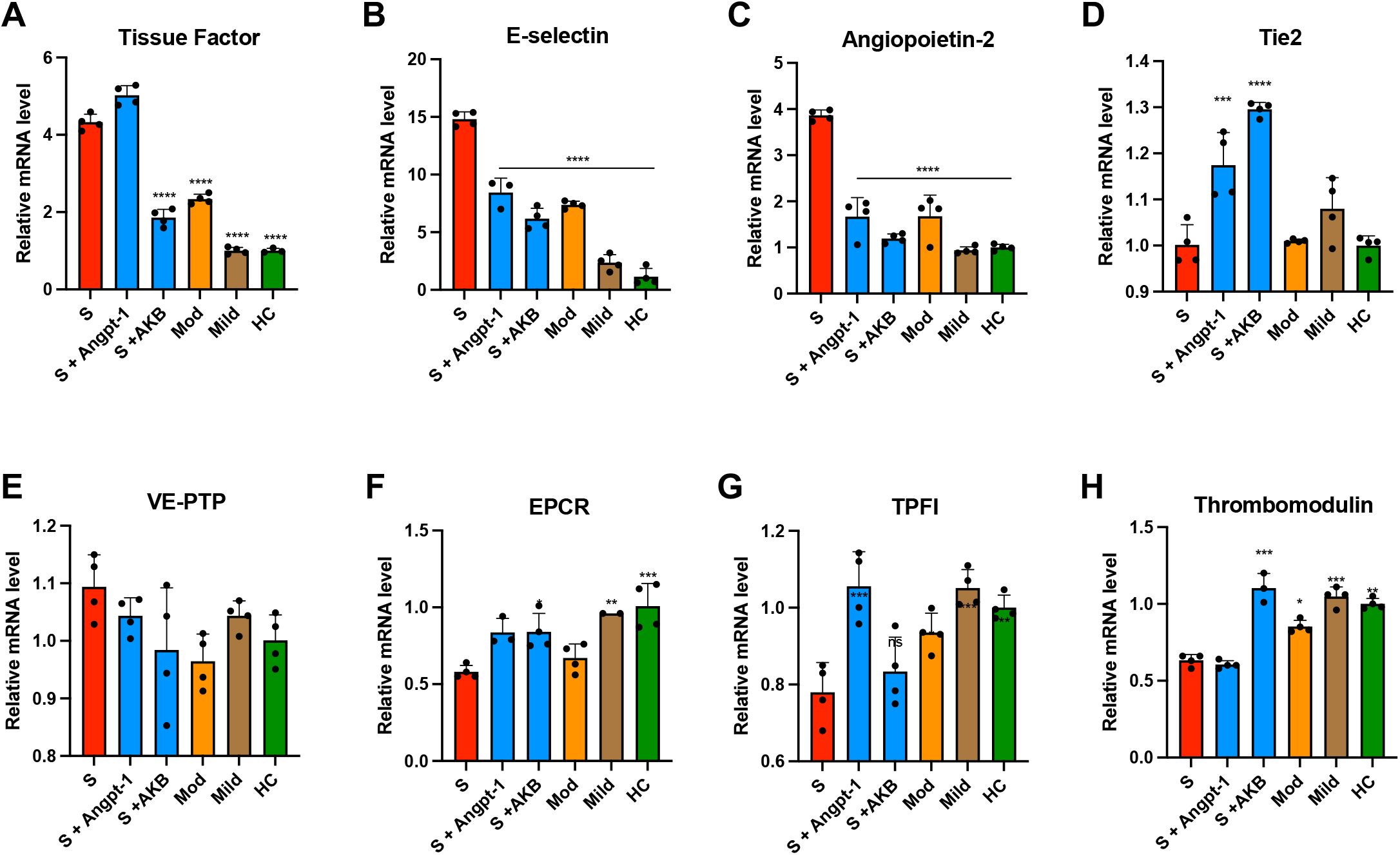
Plasma from patients with COVID-19 induces thromboinflammatory gene expression in endothelial cells. HUVECs were cultured overnight in the presence of 10% pooled plasma from patients with severe (S, ICU patients), moderate (Mod, non-ICU hospitalized patients), or mild (non-hospitalized outpatients) COVID-19 or healthy controls (HC) and analyzed for relative fold mRNA expression change of tissue factor (**A**), E-selectin (**B**), angiopoietin-2 (Angpt-2, **C**), Tie2 (**D**), vascular endothelial protein tyrosine phosphatase (VE-PTP, **E**) endothelial protein C receptor (EPCR, **F**), tissue factor pathway inhibitor (TFPI, **G**), and thrombomodulin (**H**). When indicated, cells were pretreated with Angpt-1 (300 ng/mL) or AKB-9778 (5 µM) for 30 min prior to incubation with plasma. Gene expression was normalized to that of actin and changes are shown relative to HC. Graphs represent the mean ± SD. Significance in comparison to severe (S) was determined by 1-way ANOVA using Dunnett post-test, \**P* < 0.05 \*\**P* < 0.01, \*\*\**P* < 0.001 \*\*\*\**P* < 0.0001.

We further evaluated the ability of humoral factors present in plasma from patients with COVID to promote procoagulant changes in the endothelium by measuring endothelial factor Xa and thrombin generation following exposure to COVID-19 plasma. These are the final steps immediately preceding fibrin formation and are promoted by inflamed, but not quiescent, endothelial cells. Plasma from both severe and moderate COVID-19 patients stimulated tenase and prothrombinase activity on the endothelial surface (**Figure 2**). Treatment with Tie2 activator Angpt-1 or AKB-9778 significantly attenuated the ability of plasma from severe COVID-19 to promote tenase and prothrombinase on endothelial cells. Furthermore, plasma from severe COVID-19 patients induced externalization of endothelial phosphatidylserine, an anionic phospholipid critical for assembly of coagulation enzyme complexes (**Figure 2**). Treatment with Angpt-1 or AKB-9778 also reduced PS externalization in response to severe COVID-19 plasma. These results suggest that plasma from moderate and severe COVID-19 patients promotes a prothrombotic state in endothelial cells, and that activation of Tie2 can inhibit this response.

**Figure 2.**
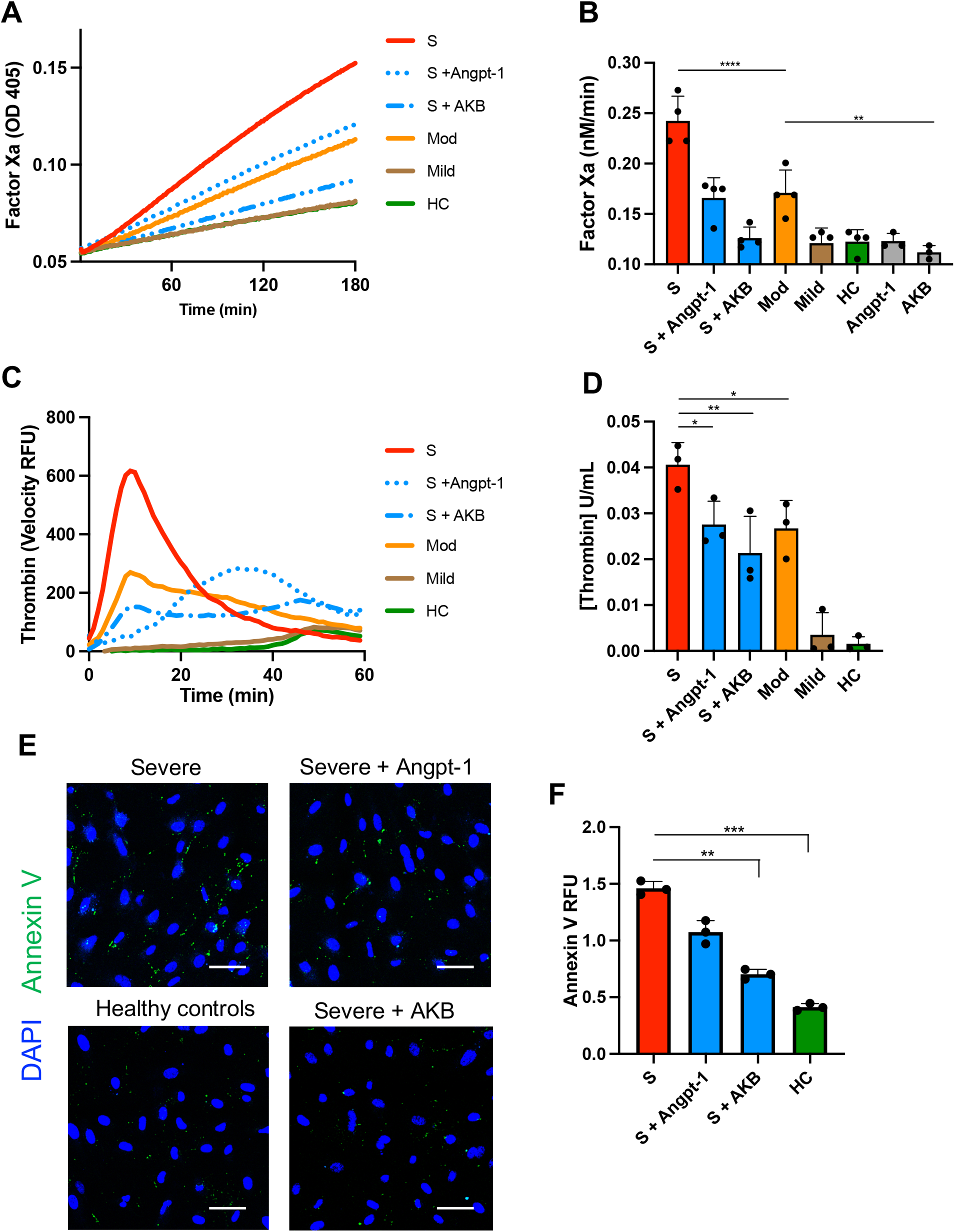
Plasma from patients with COVID-19 promotes activation of coagulation on endothelial cells. HUVECs were cultured overnight in the presence of 10% pooled plasma from patients with severe (S, ICU), moderate (Mod, non-ICU), or mild (outpatient) COVID-19 or healthy controls (HC) and analyzed for their ability to generate factor Xa (**A** and **B**) or thrombin (**C** and **D**). When indicated, cells were pretreated with Angpt-1 (300 ng/mL) or AKB-9778 (5 µM) for 30 min prior to incubation with plasma. Representative experiments are depicted as mean absorbance (405 nm) (**A**) or the first derivative of arbitrary fluorescent units as a function of time (**C**). **B** and **D**. The rate of reaction for factor Xa and thrombin were converted to nM/min and U/mL, respectively, by comparison to standard curve. **E** and **F**. Cells were stained with annexin V to assess for phosphatidylserine externalization. Total fluorescent area was quantified and normalized for number of nuclei. Graph in (**F**) represent the mean total fluorescence per 3 × 3 tile scan image at 20X magnification ± SD. Significance was determined by 1-way ANOVA using Dunnett’s post-test, \**P* < 0.05, \*\**P* < 0.01, \*\*\**P* < 0.001, \*\*\*\**P* < 0.0001.

### Increased protein markers of thrombosis in COVID-19 lung biopsies

We next sought to examine changes in expression in COVID-19 lung specimens of endothelial proteins associated with thrombotic risk. Core needle biopsies were obtained during limited autopsies performed shortly after expiration in five COVID-19 patients (see **Supplemental Appendix**). Histopathologic evaluation revealed lesions similar to those described in published autopsy series—including fibrin plugs within alveolar spaces and associated microvascular thrombi—suggesting that core needle sampling was representative of typical findings of COVID-19^32^. We compared these specimens to contemporaneously obtained normal lung tissue regions from tumor resections in non-COVID patients. Despite limited COVID-19 autopsy specimen availability, we observed increased levels of the prothrombotic endothelial protein von Willebrand factor (VWF) and decreased levels of the antithrombotic endothelial proteins EPCR and thrombomodulin (**Figure 3**).

**Figure 3.**
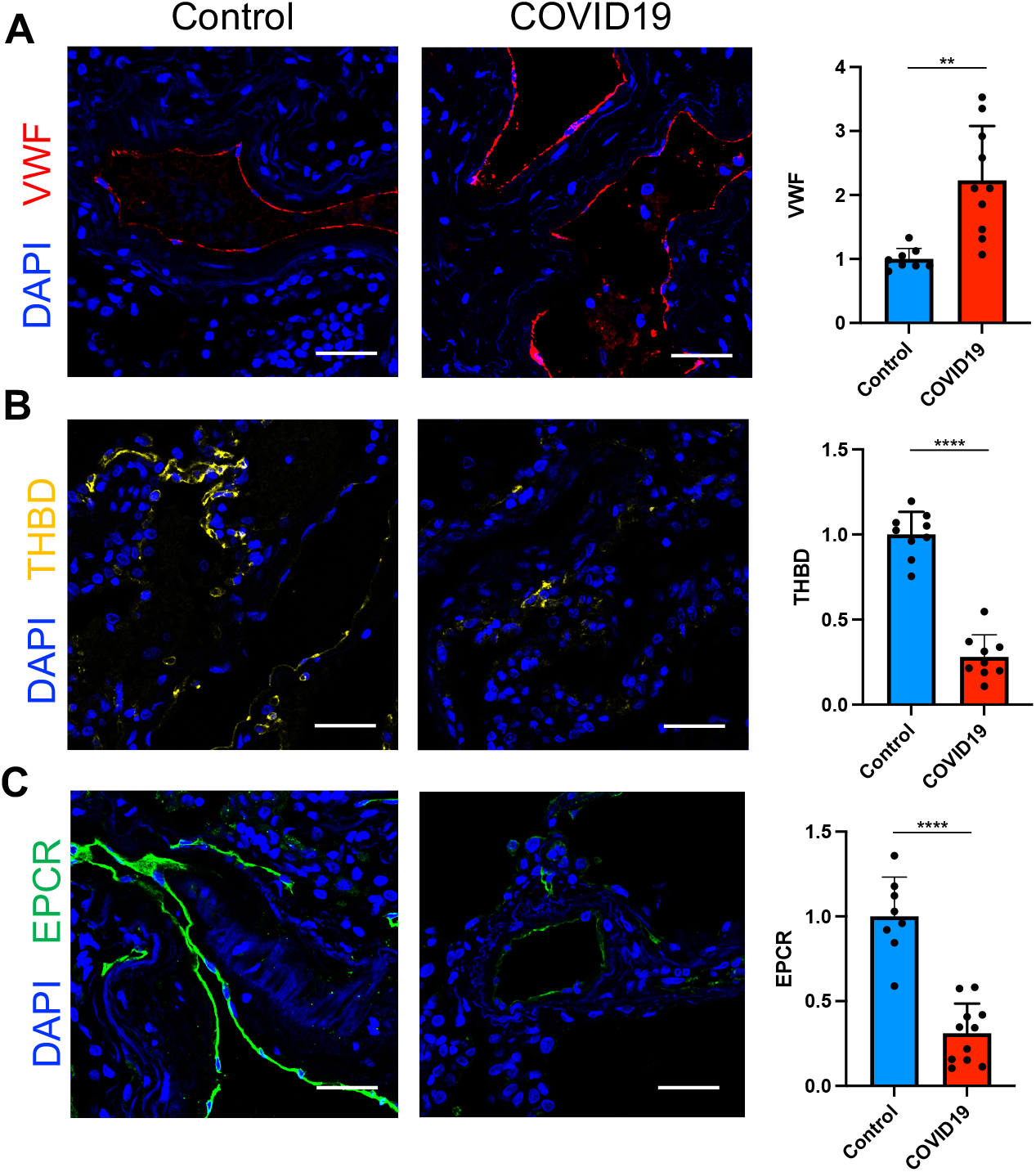
Procoagulant tissue signature of COVID-19 lung endothelium. Lung specimens were obtained during limited autopsy immediately after death in patients who died from COVID-19 (*n* = 5). Control samples (*n* = 4) were obtained from tumor-free margins of lung tumor resections and processed in identical fashion regarding timing and method of fixation. Lung specimens from COVID-19 patients demonstrated an increase in the prothrombotic endothelial protein von Willebrand Factor (VWF, **A**) and loss of the antithrombotic factors thrombomodulin (THBD, **B**) and the endothelial protein C receptor (EPCR, **C**). Two images were obtained per tissue section and mean fluorescence intensity was analyzed for each tile-scanned image and normalized to background intensity. For graphs, mean is represented by the bar with each dot as a replicate, error bars indicate SD. Significance was determined by a 2-tailed Mann-Whitney test, \*\**P* < 0.01, \*\*\*\**P* < 0.0001.

### Markers of endothelial dysfunction and thrombosis are strongly correlated with COVID-19 disease severity and survival

We measured plasma concentration of proteins associated with thrombosis and procoagulant endothelial activation and correlated these values with the degree of COVID-19 severity (**Figure 4**). Our thromboinflammtory panel included markers of general thrombotic activation, namely D-dimer and tissue factor; markers of procoagulant endothelial activation, VWF and P-selectin; and endothelial inflammation markers, E-selectin and soluble VEGFR-1. We also investigated levels of the anti-thrombotic endothelial proteins, endothelial protein C receptor (EPCR), TFPI, and thrombomodulin, which are cleaved from inflamed endothelium. Finally, given its role linking vascular inflammation and thrombosis, we investigated components of the Tie2-angiopoietin pathway, specifically Tie2-activating cytoprotective Angpt-1, Tie2-inhibiting proinflammatory Angpt-2, and soluble Tie2, which is also cleaved from endothelial cells during inflammatory states^26^.

**Figure 4.**
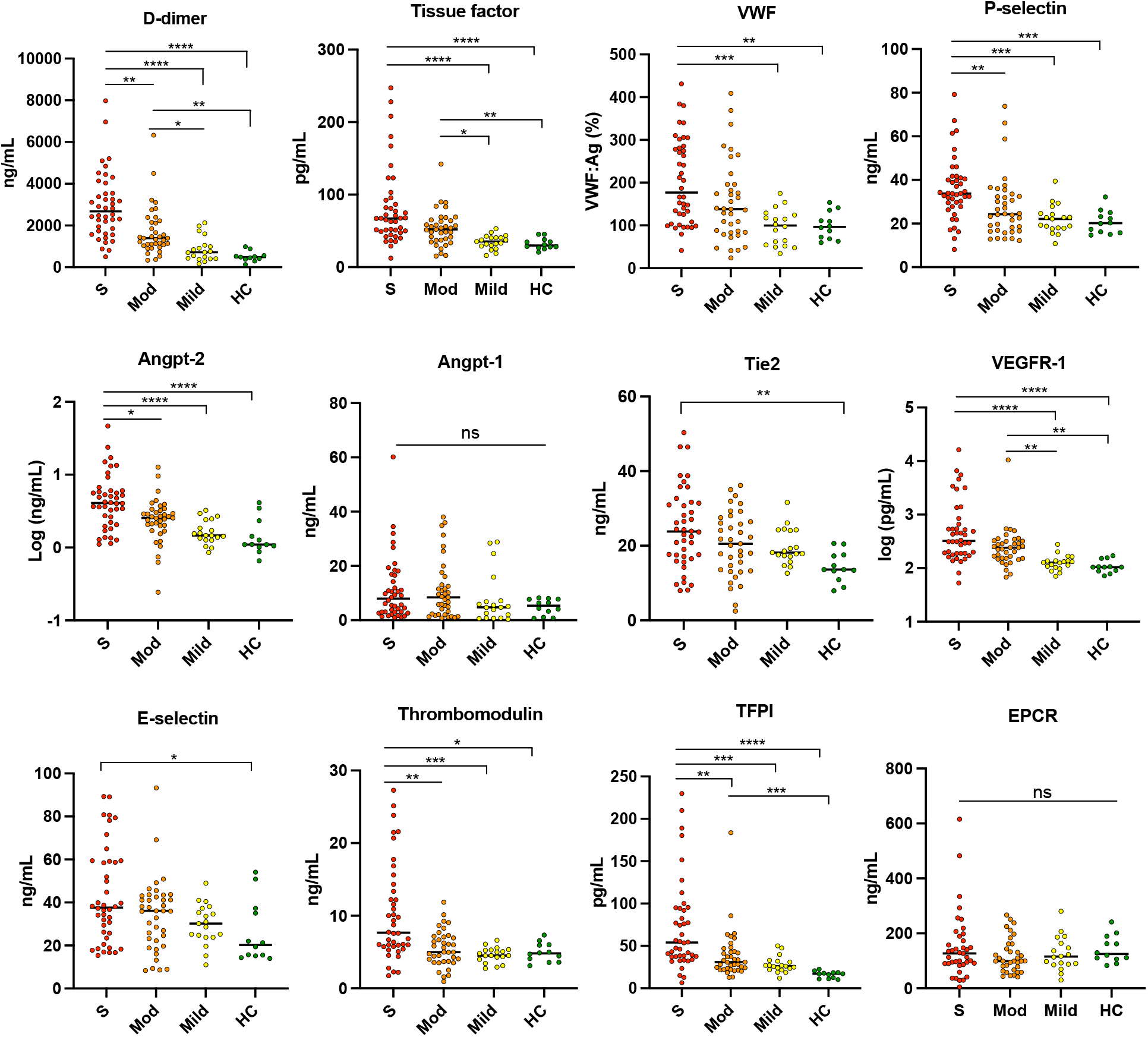
Measurement of thrombotic and endothelial markers in COVID-19 patients by disease severity. Each data point represents individual plasma measurements corresponding to disease severity. S, severe (ICU patients); Mod, moderate (hospitalized, non-ICU), Mild (COVID-19-positive outpatients); HC, healthy controls. Angpt, angiopoietin; EPCR, endothelial protein C receptor; TFPI, tissue factor pathway inhibitor; VEGFR-1, vascular endothelial growth factor receptor 1; VWF Von Willebrand Factor. Bar indicates median value. Significance among groups determined by Kruskal-Wallis with Dunn’s multiple comparison test, \**P*<0.05, \*\**P*<0.01, \*\*\**P*<0.001, \*\*\*\**P*<0.0001.

Several thromboinflammatory proteins were significantly higher in severe versus mild disease: tissue factor (*P* < 0.0001), VWF (*P* < 0.001), P-selectin (*P* < 0.001), Angpt-2 (*P* < 0.0001), VEGFR-1 (*P* < 0.0001), thrombomodulin (*P* < 0.001), and TFPI (*P* < 0.001). Angpt-2 (*P* < 0.05), along with P-selectin (*P* < 0.01), thrombomodulin (*P* < 0.01), and TFPI (*P* < 0.01) were also significantly elevated in ICU compared to non-ICU patients. There were no significant differences between groups in circulating Angpt-1 and EPCR concentrations. E-selectin and Tie2 were only significantly different between patients with severe COVID-19 versus healthy controls.

There was significant correlation among markers of prothrombotic endothelial activation. Angpt-2 levels were highly correlated with D-dimer, E-selectin, thrombomodulin, P-selectin, and VWF (**Supplemental Figure 1, top**). D-dimer correlated with P-selectin, tissue factor, thrombomodulin, VWF, and TFPI. Levels of many thromboinflammatory proteins were elevated in patients who developed acute kidney injury or died during their index hospital admission (**Supplemental Figure 2**). The relatively small number of thrombotic events, with the majority being catheter-related thrombosis, did not make it possible for us to draw meaningful conclusions regarding the association of endothelial dysfunction and clotting events in this cohort.

We performed survival analysis for selected analytes, including ones that were significantly higher among patients who died during index hospitalization. Given there were 12 deaths among 42 ICU patients, we stratified ICU patients by tertiles and analyzed survival in the top tertile of analyte concentration versus the bottom tertile. Patients in the highest tertile of Angpt-2 (Log-rank *P* = 0.048), E-selectin (Log-rank *P* = 0.019) and P-selectin (Log-rank *P* = 0.044) had worse survival compared to patients in the bottom two tertiles (**Figure 5**). These results suggest that Angpt-2 and other markers of endothelial dysfunction and thrombosis are strongly correlated with COVID-19 disease severity and implicate perturbation of the Tie2-angiopoietin pathway in this process.

**Figure 5.**
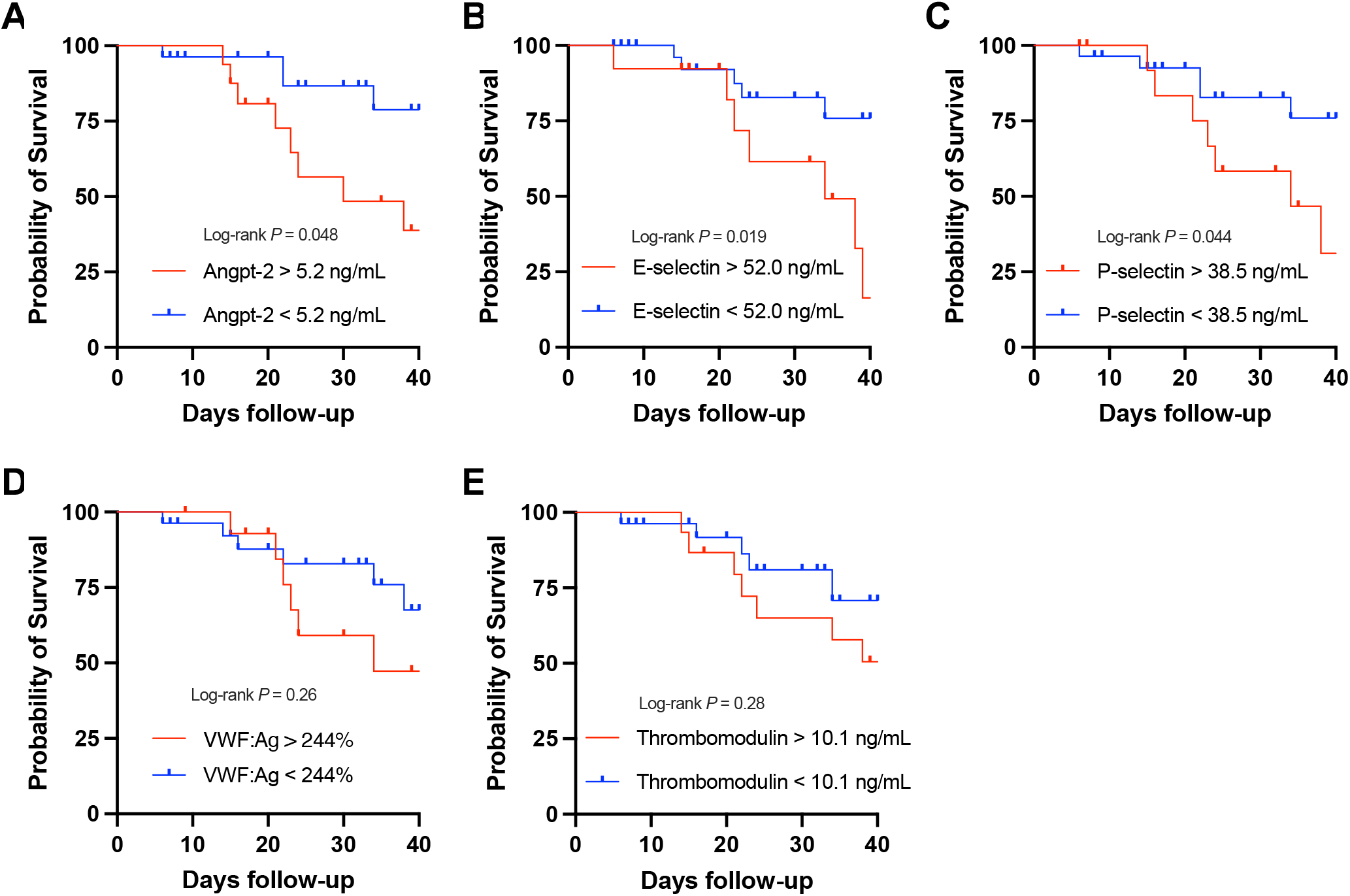
Markers of thrombotic and endothelial activation are associated with worse survival among COVID-19 patients in the ICU. Kaplan-Meier curves of survival according to the top tertile (*n* = 14) versus bottom two tertiles (*n* = 28) of the plasma concentration of indicated analytes: (A) Angiopoietin-2, (B) E-selectin, (C) P-selectin, (D) Von Willebrand factor (VWF:Ag), and (E) thrombomodulin. Significance was determined by Log-rank test.

## Discussion

To interrogate the Ang2-Tie2 system in COVID-19-mediate endothelial dysfunction, we used an *in vitro* model in which primary endothelial cells were treated with COVID-19 plasma. The effects of COVID-19 patient plasma on endothelial cells are notable in two related but distinct ways. First, there is a coordinated response such that pro-inflammatory and pro-thrombotic genes are upregulated whereas anti-inflammatory or anti-thrombotic genes are downregulated. Second, the effects of COVID humoral milieu extend to alterations of the endothelial cell surface that catalyze clot formation. Severe COVID-19 is associated with elevated levels of inflammatory cytokines, most notably TNFα, IL-6, and IL-8^1,9,10,33^, that scale with the degree of disease severity. These cytokines are known to promote transcription of prothrombotic genes such as tissue factor and selectins and to decrease platelet inhibitory nitric oxide^34,35^. Similar or even more profound cytokine elevations are associated with other critical illnesses including severe septic shock and cytokine release syndrome^9,36^, conditions also associated with profound endothelial injury and microvascular thrombosis. Loss of the constitutive endothelial anticoagulants, such as TFPI, EPCR, and thrombomodulin, is an important component of the prothrombotic endothelial transformation and may be an underappreciated driver of COVID-19 coagulopathy.

A primary function of the endothelium is to provide barrier defense to prevent excessive vessel permeability and an antithrombotic surface to promote blood circulation. Tie2 has a critical role in this process and remains activated throughout healthy adult vasculature via continuous secretion of Angpt1 from perivascular cells and platelets. As a receptor tyrosine kinase nearly exclusive to endothelial cells, Tie2 signaling promotes vascular quiescence by activating Kruppel-like factor 2 (KLF2)^37^ and inhibiting inflammatory nuclear factor-kB (NF-kB)^38^, thus promoting expression of anticoagulant genes^39^ and repressing tissue factor expression^23,40^. In our study, two unrelated approaches to Tie2 activation, direct Tie2 agonism with Angpt-1 and VE-PTP antagonism with AKB-9778, inhibit procoagulant changes in endothelial cells induced by plasma from COVID-19 patients. This result is consistent with the ability of Tie2 activation to suppress proinflammatory endothelial phenotypes in non-COVID infectious diseases such as gram-negative sepsis, anthrax, and malaria^23,41,42^. Our findings support previous work suggesting that preservation of Tie2 signaling is both *necessary* to prevent hypercoagulation and *sufficient* to normalize pathological thrombosis during systemic inflammation^23^.

VE-PTP interacts closely with Tie2 and is a natural brake on Tie2 activity. VE-PTP is highly expressed in the lung, the organ that also bears the highest concentration of Tie2. Like the other endogenous Tie2 antagonist, Angpt-2, VE-PTP is induced by pulmonary vascular stressors such as hypoxia^28^. AKB-9778 (Razuprotafib), has demonstrated beneficial activity in multiple animal models of vascular leak including LPS-induced acute lung injury^30^. Applied to cultured endothelial cells, AKB-9778 achieves ligand-independent Tie2 activation and activates Tie2 even when Angpt-1 is unable to do so during the high VE-PTP state of endothelial hypoxia. In the present study, we demonstrate that AKB-9778 strongly suppresses the procoagulant response in endothelial cells induced by COVID-19 plasma. AKB-9778 was more efficacious than Angpt-1, which may reflect the ability of VE-PTP inhibition to promote the weak agonist properties of Angpt-2^43^. Tie2 activation by AKB-9778 may therefore be an extremely effective means of dampening the thromboinflammtory state of the endothelium in COVID-19. In aggregate, our findings support clinical trials of AKB-9778 to improve pulmonary outcomes and mortality in moderate to severe COVID-19 (RESCUE, https://clinicaltrials.gov/ct2/show/NCT04511650). In addition to promoting endothelial barrier function to reduce the incidence or severity of acute respiratory disease syndrome, by lowering the procoagulant potential of the endothelium, AKB-9778 may also inhibit the microvascular thrombosis that is a hallmark of COVID-19 pathophysiology. The assays employed in this study also suggest a method to evaluate other endothelial-cell targeted therapies and assess individual-level prothrombotic risk.

While several studies have measured circulating proteins in COVID-19 patients, few to date have examined endothelial markers in lung histopathological specimens of SARS-CoV-2 infection. We attempted to examine expression of prothrombotic and antithrombotic endothelial proteins *in situ*, in lung tissue from COVID-19 autopsies. Our results demonstrate upregulation of VWF and loss of thrombomodulin and EPCR in lung tissue from COVID-19. It is therefore likely microvascular thromboses are driven at least in part by prothrombotic endothelial cell changes at the local tissue level. The profound downregulation of EPCR and thrombomodulin suggest that loss of constitutive antithrombotic function may promote clotting in COVID-19. Increased circulating thrombomodulin levels in severe COVID-19 likely reflect its cleavage from the cell surface^22^. Loss of EPCR in lung specimens was unexpected because circulating EPCR was not significantly different between groups in our cohort, even comparing critical COVID-19 disease to healthy controls (**Figure 5**). Nevertheless, the dramatic lack of EPCR antigen in COVID-19 lung specimens suggests mechanisms other than cleavage from the cell surface, such as downregulation of gene expression, may be responsible for loss of anticoagulant endothelial proteins. Indeed, plasma from patients with severe COVID-19 inhibited expression of EPCR by nearly 50% (**Figure 1**).

Our plasma vascular survey demonstrates that increasing severity of COVID-19 is associated with coagulopathy and shows a clear relationship between increasing levels of endothelial cell dysfunction and increasing clinical strata of disease severity^15,20,21^. Our data provide further support that extensive endothelial inflammation occurs in severe COVID. This study builds on prior work by demonstrating that the degree of endothelial dysfunction scales with COVID-19 severity and with procoagulant biomarkers. Specifically, markers supporting a procoagulant endothelial phenotype including Angpt-2, P-selectin, thrombomodulin, and TFPI were all elevated in severe disease compared to moderate COVID-19 in our cohort. Our data are in agreement with several lines of evidence suggesting endothelial dysfunction promotes the coagulopathy of critical illness^23^ and is a central driver of the pathobiology of severe COVID-19^8^.

Previous studies suggest that circulating Angpt-2, thrombomodulin, and VWF levels predict adverse outcomes in COVID-19^15,21,44^. We expand this list to include E-selectin and P-selectin as well. Correlation of Angpt-2, P-selectin, and VWF levels (**Fig. S1**) in our work and that of others suggest that Weibel-Palade body extrusion may be a common manifestation of COVID-19 endotheliopathy. Altogether, these findings suggest that a coordinated phenotypic shift of the endothelium contributes to adverse outcomes. Our current data add to mounting evidence that Angpt-2 is a highly sensitive and specific indicator of endothelial damage in acute illness, with COVID-19 being no exception to this trend. Extensive research has documented that Angpt-2 is a powerful biomarker for outcomes of acute respiratory distress syndrome and sepsis^27,45^, and our current findings extend prior findings implicating Angpt-2 in the coagulopathy of critical illness^23^. Indeed, data suggest that Angpt-2 levels are increased in severe COVID-19 and may predict adverse outcomes such as ICU admission^18^, acute kidney injury^46^, and survival^44^.

Our study has several limitations. Given the retrospective and non-consecutive nature of patient recruitment into the biorepositories, several biases may have inadvertently been introduced. It would have been useful to have readouts of gene expression and coagulation assays of individual plasmas correlated with markers detected *in vivo* or whether these functional assays provided additional predictive clinical information. Given the limited amount of plasma supply however, it was necessary to pool the individual plasmas. The amount of autopsy specimens were limited, which impaired our ability to perform comprehensive histopathological assessment of multiple endothelial markers. We cannot rule out that differences in specimen preparation affected antibody staining. That samples were not collected uniformly early upon hospital or ICU admission limits the applicability of our conclusions regarding endothelial biomarkers and clinical endpoints.

In conclusion, the present study supports the concept that moderate and severe COVID-19 are driven at least in part by procoagulant endothelial cell dysfunction, the degree of which increases in parallel with COVID-19 disease severity. Elevated Angpt-2 levels may potentiate endothelial cell dysfunction through inhibition of antithrombotic Tie2 signaling. Activation of Tie2 through two independent mechanisms corrects the prothrombotic changes in endothelial cells induced by plasma from COVID-19 patients. Our findings support further investigation into the role of Tie2-angiopoietin in SARS-CoV-2 infection and encourage clinical trials to evaluate the efficacy of Tie2-activating therapy for treatment of more severe forms of COVID-19.

## Materials and Methods

### Patient cohort and sample collection

Subjects were recruited between April and June 2020, during the height of the COVID-19 surge, at Beth Israel Deaconess Hospital (BIDMC) by the BIDMC COVID-19 Data and Tissue Repository and at Massachusetts General Hospital (MGH) by the Massachusetts Consortium on Pathogen Readiness. Patients were recruited who had PCR-confirmed SARS-CoV-2 either in the hospital or ambulatory setting. All enrolled patients provided either written and/or verbal informed consent, participation was entirely voluntary, and the repository studies were approved by the internal review board (IRB) at their respective institutions (BIDMC IRB 2020P000361 and MGH IRB 2020P000804). Blood samples were collected into ethylenediamine tetraacetic acid (EDTA) tubes and spun for 15 min at 2600 rpm according to standard protocol. Plasma was aliquoted into 1.5-mL cryovials and stored at -80C, which was subsequently thawed and further aliquoted for this study. Demographics, laboratory values, and clinical endpoints were assessed through review of the patients’ electronic medical record. Acute kidney injury was defined as a rise in serum creatinine of > 0.3 mg/dL within 48 hours or an increase to > 1.5 times baseline over 7 days, and patients with end-stage renal disease were excluded from this analysis. Thrombotic events were reviewed and included deep vein thrombosis, pulmonary embolism, catheter-related thrombosis, or clinically documented thrombosis at another location.

### Plasma biomarker measurements

Plasma was thawed on ice, then centrifuged for 5 min to pellet any debris. Concentrations of the following proteins were measured using a Luminex Human Premixed Multi-Analyte Kit (R&D Systems) for the following analytes: Angpt-1, Angpt-2, D-dimer, E-selectin, P-selectin, thrombomodulin, Tie2, Tissue factor, VWF, and VEGFR1, according to manufacturer’s protocol. Samples were diluted 1:2 and run on a MAGPIX system (Millipore Sigma) which was preprogrammed according to kit specifications. For EPCR, plasma samples were analyzed by ELISA (Diagnostica Stago) at 1:51 dilution according to manufacturer’s protocol. For TFPI, plasma samples were analyzed by ELISA (R&D systems) at a 1:100 dilution according to manufacturer’s protocol.

### Human lung tissue isolation and immunofluorescence microscopy

Core needle biopsies were obtained during limited autopsy of COVID-19 patients within 3 h of patient expiration. For controls, surgical specimens from patients undergoing lung tumor resections were analyzed by a pathologist, and lung tissue within the tumor-free margins was isolated. These controls were selected due to the similar nature in which the lung tissue was processed compared to standard autopsy where there is a longer delay before tissue is placed in fixative. Tissue was washed and fixed in freshly prepared paraformaldehyde 4% for 24 h and transferred to 70% ethanol. Samples were embedded in paraffin within 7-10 days and cut into 6-µm-thick sections for staining. Primary antibodies used for immunofluorescence microscopy are as follows: anti-VWF (clone IIIE2.34, Millipore Sigma), anti-thrombomodulin (clone 141C01, Thermo Fischer Scientific), and anti-EPCR (clone LMR-42, Thermo Fisher Scientific). Images were obtained using a Zeiss LSM 880 upright laser scanning confocal microscope in 3 × 3 tile-scan mode with a Plan-Apochromat 40X/1.3 Oil DIC M27 objective.

### Endothelial cell culture

Human umbilical vein endothelial cells (HUVEC, pooled donor, Lonza) were grown on collagen-coated culture plates and maintained in endothelial cell growth basal media (EBM-2, Lonza), containing 2% fetal bovine serum (FBS) and contents of the EGM-2 SingleQuots growth factor supplement kit. Cells from passages 3-5 were used for experiments. For studies on endothelial cell monolayers, plasma from severe, moderate, mild, and healthy controls were pooled. HUVECs were grown to confluency in 96-well plates and incubated overnight with 10% pooled patient plasma and H-Gly-Pro-Arg-Pro-OH (GPRP, 5 mM, Cayman Chemical) in the presence of complete growth media. Supplementation of rAngpt-1 (300 ng/mL, R&D systems,) or AKB-9778 (5 µM, a gift from Aerpio pharmaceuticals) was performed 30 min prior to addition of patient plasma. All endothelial-based studies were performed in a dedicated tissue-culture room designated specifically for work with COVID-19 biospecimens.

### Gene expression analysis

Confluent HUVECs were treated with pooled patient plasma as described above and gene expression was determined using a 2-step Cell-to-Ct Taqman kit (Thermo Fisher Scientific). The following gene expression probes (Taqman, Thermo Fisher Scientific) were used:

Hs00174057_m1 (*SELE*), Hs00264920_s1 (*THBD*), Hs00409207_m1 (*TFPI*), Hs00197387_m1 (*PROCR*), Hs01076029_m1 (*F3*), Hs00169867_m1 (*ANGPT2*), Hs00176096_m1 (*TEK*), Hs00160781_m1 (*PTPRB*), Hs00194899_m1 (*ACTB*). Quantitative reverse transcriptase polymerase chain reaction (qRT-PCR) was performed in technical duplicate for each biologic sample using a QuantStudio 6 Flex real-time PCR system. Gene expression was compared to *ACTB* control and normalized to cells treated with healthy control plasma using the Δ1Δ1Ct method.

### Factor Xa and thrombin generation assays

HUVECs treated as above were equilibrated at room temperature and treated with calcium inophore A23187 (2 µM, Sigma) for 15 min to promote tissue factor activation. Cells were washed twice with HEPES-buffered saline containing 1% fatty acid-free bovine serum albumin (HBS-BSA, pH 7.4). For Factor Xa generation experiments, cells were incubated with HBS-BSA containing factor X (125 nM, Haematologic Technologies, Factor VIIa (0.6 nM, Haematologic Technologies), and chromogenic substrate biophen-CS11(22) (150 µM, Anaira Diagnostica). Absorbance at 405 nm was measured every minute for 3 h on an xMark Spectrophotometer (Bio-Rad). Maximal reaction velocity was converted to FXa nM/min based on standard curve analysis of FXa (Haematologic Technologies) serial dilutions. For thrombin generation experiments, cells were washed twice with HBS-BSA and incubated in 80 µL HBS plus 20 µL pooled human plasma (George King Bio-Medical) to supply coagulation factors and 5 mM GPRP. Thrombin generation was measured using the fluorogenic substrate Boc-L-FPR-ANSNH-C2H5 (SN-20, Haematologic technologies). Fluorescence (excitation 352 nm / emission 470 nm) was measured every minute for 1 h using the Synergy HTX plate reader (BioTek). First derivative of thrombin generation curves were compared with a standard curve of thrombin to determine thrombin generation in U/mL. All biologic replicates were performed in technical triplicate.

### Phosphatidylserine Externalization

HUVECs were grown to confluence in glass chamber slides and incubated with pooled patient plasma as described above. Cells were washed with 10 mM HEPES buffer (pH 7.4) containing 140 mM NaCl, 2.5 mM CaCl2, and 2% FBS (annexin V binding buffer) and stained with annexin V-Alexa Fluor 488 (Thermo Fischer Scientific) at a 1:50 dilution and Zombie Red viability dye (BioLegend) at a 1:1000 dilution for 15 minutes at room temperature in annexin V binding buffer. Cells were washed and fixed in paraformaldehyde 4% / annexin V binding buffer for 7 minutes. Cells were washed 3 times and mounted with DAPI. Images were obtained using a Zeiss LSM 880 upright laser scanning confocal microscope in 3 × 3 tile-scan mode with a Plan-Apochromat 20X/0.8 M27 objective.

### Image analysis

Fluorescent images were analyzed using Image J software (NIH). For phosphatidylserine externalization, annexin V staining was thresholded and total fluorescent area was normalized to the number of nuclei. For immunofluorescence microscopy of lung sections, fluorescence intensity was quantified per tissue area, after measuring and subtracting background signal from each image.

### Statistics

Tests of normality were performed using the Anderson-Darling and D’Agonstino-Pearson method. Statistical significance for binary comparisons of continuous variables were assessed by unpaired two-tailed Student’s *t* test unless the data did not demonstrate normality, in which case differences between groups were analyzed by Mann-Whitney *U* test. For comparison of continuous variables across multiple groups, all of which did not pass the test of normality, with the exception of Tie-2, the Kruskal-Wallis test with Dunn’s post-hoc for multiple comparisons was performed. Correlation matrix analysis was performed using two-tailed nonparametric Spearman correlation. Survival analysis for ICU-patients was performed by segregating patients into the top tertile versus bottom two tertiles for each analyte. Kaplan-Meier analysis was performed, and survival was compared for the top tertile versus bottom two tertiles using the Mantel-Cox log-rank test. Comparison of categorical variables were performed using Fisher’s exact test. All statistical analysis was performed using GraphPad Prism (version 9.0; GraphPad Software, San Diego, CA). *P* values of less than 0.05 were considered significant.

### Study approval

This study measuring biomarkers and associated clinical data in patients enrolled in the BIDMC COVID-19 Data and Tissue Repository was approved by the BIDMC IRB (protocols 2020P000621). For all COVID-19 autopsy studies, patients were consented for limited autopsies by a pathologist during a witnessed phone call immediately after death of the patient. Research using autopsy tissue was approved by the BIDMC institutional review board (IRB 2020P000525). A HIPPA waiver was granted to access the medical records of the patients undergoing autopsy (IRB 2020P00412).

## Data Availability

There are no datasets to deposit. Other data will be made available upon request via correspondence with the authors.

## Acknowledgements

The authors would like to thank Jonathan Hecht for directing the COVID-19 autopsy program, including the acquisition and processing tissue specimens, assembly of pathology reports and obtaining appropriate control specimens. We greatly appreciate Michelle Hacker, Jonathan Li, and Michael Seaman’s support with the COVID-19 biorepositories. We thank Christiane Ferran, Cleide Angolano, and Johannes Schlondorff for their assistance in establishing the BSL2+ facility for sample processing. We are grateful to Shulin Lu for assistance with Luminex assays. We appreciate Lay-Hong Ang, Anikit Gad and the BIDMC Histology and Confocal Imaging Core for assistance with immunofluorescence staining. We would like to thank Karla Pollick and Stephanie Li from the BIDMC InSIGHT Core for help with clinical data collection. The authors acknowledge the following groups and individuals: the CORE Steering Committee – Laurie Farrell, Aarti Asnani, Elias Baedorf-Kassis, Valerie Banner-Goodspeed, Somnath Bose, Daniel Katz, Douglas Krakower, Michelle Lai, Anica Law, Jaymin Patel, Nathan Shapiro, Gyongyi Szabo, Jeffery Zwicker, and the numerous research assistants and fellows who contributed to data collection. The authors thank the participants and their families, the frontline healthcare providers, and the laboratory staff of the BIDMC COVID-19 Biorepository and MGH/MassCPR COVID biorepository (see **Supplemental Acknowledgements**). We also are grateful to the Center for Virology and Vaccine Research, the Harvard Catalyst Clinical Research Center, the BIDMC Department of Obstetrics and Gynecology, the Cardiovascular Institute at BIDMC, and the office of the BIDMC Chief Academic Officer for enrollment, collection, and processing samples for the BIDMC COVID-19 Biorepository.

## Author contributions

AAS, RF, and SMP conceived the study. AAS performed endothelial cell studies, immunofluorescence studies, measured analytes in plasma from COVID-19 patients, and analyzed biologic and clinical data. SMC performed endothelial cell culture and reagent preparation. GPH collected, organized and analyzed clinical data. ZJM, KDS, VB, and JQ collected clinical data. AYC, DN, DHB, REG, and XGY oversaw collection of biorepository specimens and clinical data from the COVID-19 cohort. KP and AJN contributed new reagents and analytic tools. AAS, RF, and SMP wrote the manuscript with input and approval from all authors.

