## Supplemental data and appendix for "Tie2 activation protects against prothrombotic endothelial dysfunction in COVID-19"

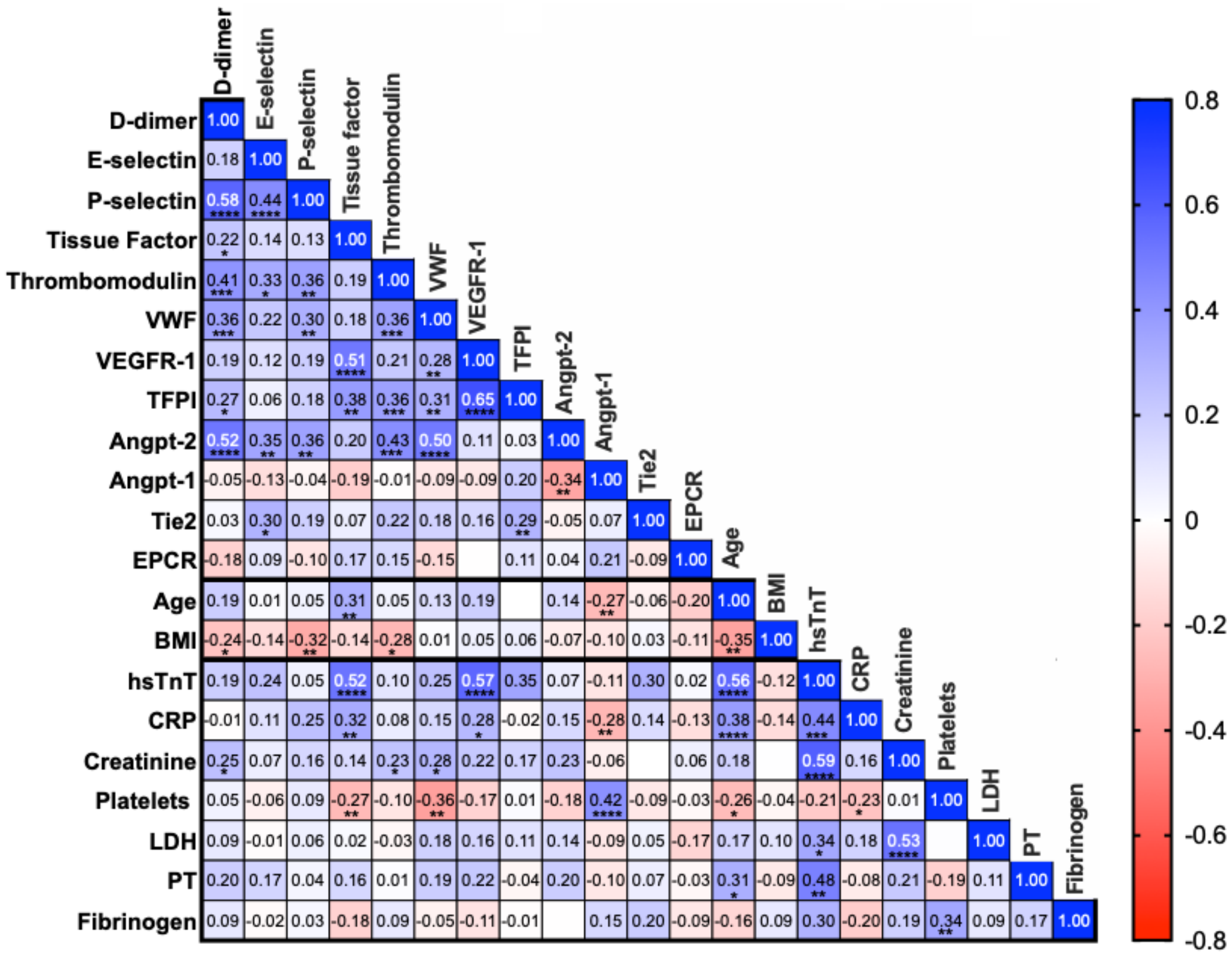


**Supplemental Figure 1. Correlation matrix of measured analytes among hospitalized COVID-19 patients.** Spearman rank correlations were performed for markers of endothelial and thrombotic activation, demographics, and clinical metrics. Positive correlations are indicated in blue and negative correlations indicated in red. Numbers within the square indicate r values. Angpt, angiopoietin; BMI, body mass index; CRP, C-reactive protein; EPCR, endothelial protein C receptor; hsTnT, high-sensitivity troponin, LDH, lactate dehydrogenase; PT, prothrombin time; TFPI, tissue factor pathway inhibitor; VEGFR1, vascular endothelial growth factor receptor 1; VWF, Von Willebrand Factor, *$P$<0.05, **$P$<0.01, ***$P$<0.001, ****$P$<0.0001.

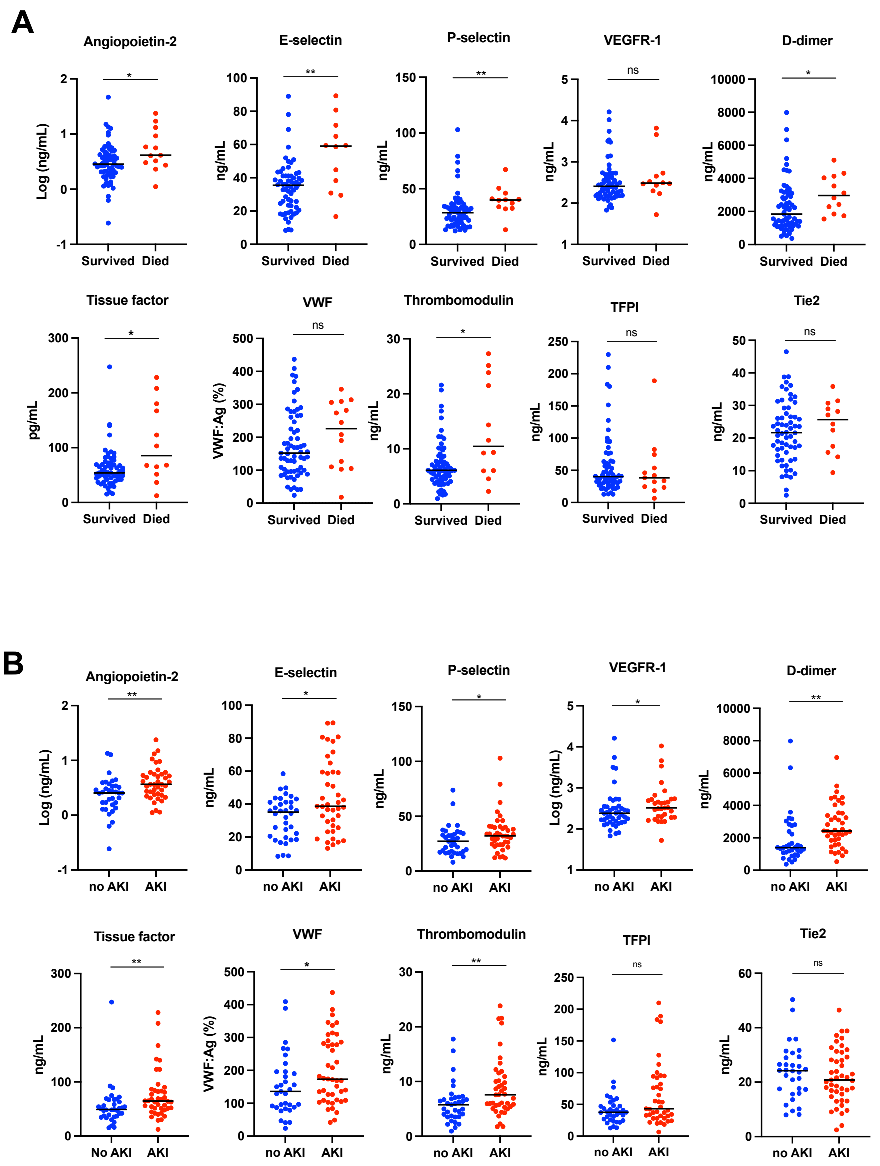


**Supplemental Figure 2. Endothelial and thrombotic activation and clinical endpoints among patients hospitalized with COVID-19.** Plasma samples obtained from patients hospitalized with COVID-19 and values of measured proteins are shown for patients according to death (**A**) or development of acute kidney injury (**B**). Acute kidney injury (AKI) was defined as a rise in serum creatinine of ≥ 0.3 mg/dL or ≥ 1.5 times baseline. TFPI, tissue factor pathway inhibitor; VEGFR-1, vascular endothelial growth factor receptor 1; VWF, Von Willebrand Factor. Bar indicates median value. Significance was determined by a 2-tailed Mann-Whitney test. *$P$<0.05, **$P$<0.01,

**Supplemental appendix**

**COVID-19 Autopsy specimens**

Three characteristic findings were apparent in the lung specimens isolated from COVID-19 autopsies—1) alveolar spaces with pulmonary edema and fresh hemorrhage, without significant interstitial inflammation 2) chronic interstitial inflammation and edema, with type II pneumocyte hyperplasia and multinucleation, and 3) diffuse alveolar damage with organizing fibrin in alveolar spaces and associated microvascular thrombi[32]

Patient 1 (10 day admission) presented with low-grade fever and malaise, and initially a negative chest x-ray without pulmonary symptoms. Labs showed lymphopenia and elevated inflammatory markers. He had profound hypotension and progressive decline in strength and mental status. He was transitioned to comfort measures and died on day 10. Autopsy findings showed lungs with patchy pulmonary edema and acute alveolar hemorrhage.

Patient 2 (16 day admission) presented with fever, hypoxia, and respiratory distress requiring oxygen supplementation and metabolic acidosis. After response to fluid resuscitation, she tested positive for COVID-19 by swab and was discharged to in-patient hospice where her respiratory status declined until death. Autopsy findings matched the clinical picture of lung injury. The lungs showed diffuse alveolar damage and microvascular thrombosis. In less involved areas, sections showed a prominent septal lymphocytic infiltrate and type-II pneumocyte hyperplasia with multinucleated forms.

Patient 3 (40 day admission) presented with 10 days of fever, chills, and dry cough as well as nausea/vomiting and diarrhea, followed by worsening dyspnea. At presentation she had bilateral lung field infiltrates on x-ray, lymphopenia and rapidly progressing hypoxia with positive COVID-19 test. She was intubated and required pressor support to treat hypotension. She required mechanical ventilation and hemodialysis for acute kidney injury. At autopsy, the lungs with prominent interstitial lymphocytic infiltrate and abundant alveolar macrophages likely related to recent pneumonias.

Case 4 (15 day admission) presented with 2 days of shortness of breath, cough and fatigue. On arrival she was hypoxemic, and chest x-ray demonstrated multifocal pneumonia and COVID testing was positive. She was intubated on day 2 with persistent hypoxemia and she developed acute respiratory distress syndrome (ARDS) and persistent shock requiring pressor support and died. At autopsy, her lungs showed patchy pulmonary edema and acute alveolar hemorrhage with prominent interstitial lymphocytic pneumonia with type-II pneumocyte hyperplasia and abundant alveolar macrophages and fibrin deposition in the microvasculature.

Case 5 (31 day admission) presented with 4 days of fever and malaise and was admitted for COVID-19-positive pneumonia requiring intubation. He required vasopressor support throughout his admission. He developed renal failure requiring hemodialysis. At autopsy, his lungs showed diffuse alveolar damage with associated microvascular thrombosis and organizing alveolar hemorrhage which correspond to diffuse radiographic infiltrates seen on daily chest x-rays.

**Supplementary Appendix Table 1.** Clinical characteristics and laboratory values for patients included in the cohort. Highest value during hospitalization is listed. (HTN – hypertension, ESRD – end stage renal disease, PE – pulmonary emboli, DM2 – diabetes mellitus type 2, TIA – transient ischemic attack, CKD – chronic kidney disease, AFib – atrial fibrillation).

| | Age range / gender | Hospital admission length | Intubated | Comorbidities | D- Dimer | cTropnT |
|---|---|---|---|---|---|---|
| **1** | 85-90, M | 10 | no | ESRD on dialysis, HTN | 1174 | 0.17 |
| **2** | 80-85, F | 16 | no | HTN, DM2, TIA | >21600 | 0.08 |
| **3** | 55-60, F | 40 | yes | HTN, DM2, obesity, mild CKD | >21600 | 0.25 |
| **4** | 75-80, F | 15 | no | ESRD on dialysis, HTN | >21600 | 0.23 |
| **5** | 70-75, M | 31 | yes | HTN, AFib | 20937 | 1.35 |

### Supplemental acknowledgments

**MGH COVID-19 Collection & Processing Team Participants**

**Collection Team:** Kendall Lavin-Parsons[1], Blair Parry[1], Brendan Lilley[1], Carl Lodenstein1, Brenna McKaig[1], Nicole Charland[1], Hargun Khanna[1], Justin Margolin1, Edward DeMers[6], Kelly Judge[6], Bruce D. Walker[6], Peggy Lai[6], Musie S. Ghebremichael[6]

**Processing Team:** Anna Gonye2, Irena Gushterova2, Tom Lasalle2, Nihaarika Sharma2, Brian C. Russo[3], Maricarmen Rojas-Lopez[3], Moshe Sade-Feldman[4], Kasidet Manakongtreecheep[4], Jessica Tantivit[4], Molly Fisher Thomas[4]

**Massachusetts Consortium on Pathogen Readiness:** Betelihem A. Abayneh[5], Patrick Allen[5], Diane Antille[5], Katrina Armstrong[5], Siobhan Boyce[5], Joan Braley[5], Karen Branch[5], Katherine Broderick[5], Julia Carney[5], Andrew Chan[5], Susan Davidson[5], Michael Dougan[5], David Drew[5], Ashley Elliman[5], Keith Flaherty[5], Jeanne Flannery[5], Pamela Forde[5], Elise Gettings[5], Amanda Griffin[5], Sheila Grimmel[5], Kathleen Grinke[5], Kathryn Hall[5], Meg Healy[5], Deborah Henault[5], Grace Holland[5], Chantal Kayitesi[5], Vlasta LaValle[5], Yuting Lu[5], Sarah Luthern[5], Jordan Marchewka (Schneider)[5], Brittani Martino[5], Roseann McNamara[5], Christian Nambu[5], Susan Nelson[5], Marjorie Noone[5], Christine Ommerborn[5], Lois Chris Pacheco[5], Nicole Phan[5], Falisha A. Porto[5], Edward Ryan[5], Kathleen Selleck[5], Sue Slaughenhaupt[5], Kimberly Smith Sheppard[5], Elizabeth Suschana[5], Vivine Wilson[5], Galit Alter[6], Alejandro Balazs[6], Julia Bals[6], Max Barbash[6], Yannic Bartsch6, Julie Boucau[6], Josh Chevalier6, Fatema Chowdhury[6], Kevin Einkauf6, Jon Fallon[6], Liz Fedirko[6], Kelsey Finn[6], Pilar Garcia-Broncano[6], Ciputra Hartana[6], Chenyang Jiang[6], Paulina Kaplonek[6], Marshall Karpell[6], Evan C. Lam[6], Kristina Lefteri[6], Xiaodong Lian[6], Mathias Lichterfeld[6], Daniel Lingwood[6], Hang Liu[6], Jinqing Liu[6], Natasha Ly[6], Ashlin Michell[6], Ilan Millstrom[6], Noah Miranda[6], Claire O'Callaghan[6], Matthew Osborn[6], Shiv Pillai[6], Yelizaveta Rassadkina[6], Alexandra Reissis[6], Francis Ruzicka[6], Kyra Seiger[6], Libera Sessa[6], Christianne Sharr[6], Sally Shin[6], Nishant Singh[6], Weiwei Sun[6], Xiaoming Sun[6], Hannah Ticheli[6], Alicja Trocha-Piechocka[6], Daniel Worrall[6], Alex Zhu[6], George Daley[7], David Golan[7], Howard Heller[7], Arlene Sharpe[7], Nikolaus sJilg[8], Alex Rosenthal[8], Colline Wong[8]

[1]Department of Emergency Medicine, Massachusetts General Hospital, Boston, MA, USA.
[2]Massachusetts General Hospital Cancer Center, Boston, MA, USA.
[3]Division of Infectious Diseases, Department of Medicine, Massachusetts General Hospital, Boston, MA, USA.
[4]Massachusetts General Hospital Center for Immunology and Inflammatory Diseases, Boston, MA, USA.
[5]Massachusetts General Hospital, Boston, MA, USA.
[6]Ragon Institute of MGH, MIT and Harvard, Cambridge, MA, USA.
[7]Harvard Medical School, Boston, MA, USA.
[8]Brigham and Women's Hospital, Boston, MA, USA.
